# Associations between Multiple Early Risk Factors and Cognitive Functioning at Time of Diagnosis in Children, Adolescents, and Adults with First-Episode Psychosis

**DOI:** 10.64898/2026.05.11.26352862

**Authors:** CK Lemvigh, WT Syeda, KMS Ambrosen, JRM Jepsen, MØ Nielsen, J Rydkjær, KB Bojesen, NK Andersen, C Pantelis, AK Pagsberg, BY Glenthøj, M Osler, B Fagerlund, BH Ebdrup

## Abstract

**Background:** Schizophrenia is associated with widespread cognitive impairments. Several early risk factors for schizophrenia have been identified, and some studies suggest associations between these early risk factors and cognition, yet the literature is sparse in psychosis.

**Methods:** Clinical cohorts of child/adolescent and adult patients with first-episode psychosis (FEP) and healthy controls (HC) were linked with register-based information (N=276). Gestational age, Apgar scores, birth weight and length, parental age, and season of birth were extracted from the Danish medical birth registry. Cognition was assessed at time of diagnosis using BACS, CANTAB, and WAIS-III/WISC-IV. Missing data was imputed using MICE. Canonical correlation analysis (CCA) was used to examine patterns of associations. Post hoc analyses explored group differences according to diagnosis, age, and sex.

**Results:** CCA resulted in two significant patterns of associations. CCA1 was stable across imputations (r=0.44, *p*=.036, p_min_= .017, p_max_= .055), and constituted by a risk profile of lower maternal age, lower birth length, being small for gestational age, and lower birth weight and a cognitive profile of lower estimated IQ and poorer working memory, mental flexibility, processing speed, verbal fluency, and motor latency. The pattern was more expressed in FEP compared to HC and in adults compared to children. CCA2 was more unstable across imputations (r=0.38, *p*=.033, p_min_=.003, p_max_=.168) and constituted by a broad pattern of minor loadings.

**Conclusion:** Cognitive functioning later in life is associated with multiple early risk factors, underscoring the complexity of developmental processes and the importance of the early developmental context in shaping cognitive trajectories.

## INTRODUCTION

Patients with schizophrenia present with widespread cognitive deficits already from their first psychotic episode^1,2^. Some evidence suggests that these cognitive deficits are neurodevelopmental in origin, with individuals who later develop a psychotic disorder showing an increasing lag in cognitive development compared to non-psychiatric peers^3,4^. The diagnosis of schizophrenia is typically made in early adulthood^5,6^, however, some patients experience their first psychotic episode during childhood or adolescence, referred to as early-onset psychosis^7^. Patients with early-onset psychosis also experience a broad range of cognitive deficits^8,9^.

Twin studies have demonstrated that most cognitive functions are heritable, although to a varying degree depending on the specific domain and tests used. Heritability estimates range from 20%-40% for executive functions, 21%-64% for visual and verbal memory and working memory, 28%-74% for attention and processing speed to 52%-86% for measures of intelligence (IQ)^10–12^. Moreover, schizophrenia itself is a highly heritable disorder^13^ and studies have reported a genetic association between cognition and schizophrenia liability, indicating a partially overlapping etiology^11,12,14^. In addition, numerous early risk factors for schizophrenia have been identified including, but not limited to, low birth weight, obstetric complications, advanced paternal age and winter/spring birth^15,16^. How these early risk factors influence cognitive development is less clear and the literature in patient samples is sparse.

In healthy individuals, meta-analyses have shown a positive association between birth weight and IQ in adolescence and adulthood^17–19^. For example, a large perinatal cohort from Denmark followed until the age of 50 years revealed stable associations between birth weight and IQ levels across three timepoints^20^. Moreover, a recent study of 109,481 children from eight European birth cohorts showed that preterm birth and being small for the gestational age (SGA) were associated with lower non-verbal IQ^21^. The association between preterm birth or SGA and lower IQ has been replicated in different countries^19,22,23^.

An inverse U-shaped relationship between paternal age and IQ in the offspring has also been reported, with lower IQ in children of both the youngest and oldest fathers^24^. However, some evidence suggests that this association may be explained by maternal age, parental education or birth order^25^. A meta-analysis reported more cognitive and learning problems in adolescents of young mothers (<21 years old)^26^. Moreover, in a large study covering three birth cohorts the association between advanced maternal age (age 35-39 years) and cognition in the offspring changed from negative in older cohorts to positive in the newest (born between 2000-2002), which may be explained by changing parental characteristics with higher maternal age being associated with more advantaged socioeconomical background^27^. Only a handful of studies have examined the relationship between winter birth and cognition^28–32^, and findings are mixed with reports of both better and lower cognitive abilities as well as no associations.

In patients with schizophrenia, a meta-analysis reported lower verbal memory and working memory in patients with a history of obstetric complications compared to those without^33^. Moreover, in a small study of patients with early-onset psychosis, lower gestational age and Apgar scores were associated with lower executive functioning^34^. Finally, some evidence has indicated an association between birth weight and cognition in patients with schizophrenia^35,36^. Interestingly, in this population, both low and high birth weight have been linked to cognitive deficits^35,36^.

In sum, the existing literature suggests that early risk factors for schizophrenia may be associated with impairments across several cognitive domains, however, the sparse literature suggests complex associations. Against this background, we aimed to examine the relationship between early risk factors for schizophrenia obtained at birth and a broad range of cognitive functions assessed at the time of diagnosis in patients with first-episode psychosis across various age groups and matched healthy controls (HC). In post hoc analyses, we explored potential group differences in these associations between patients and HC, children and adults, females and males. A better understanding of the patterns of associations between early risk factors and cognitive functioning is important in terms of early identification and prevention strategies.

## METHODS

Four clinical cohorts of child, adolescent, and adult patients with first-episode psychosis (FEP) and matched healthy controls (HC) were recruited for collaborative studies between child- and adolescent psychiatry and the Center for Clinical Intervention and Neuropsychiatric Schizophrenia Research from 2008–2017 (PECANS I^37^, PECANS II^38^, ATT^39^ & DBUP^40^). The four cohorts used a partly overlapping comprehensive clinical and cognitive battery. Some cognitive data pertaining to the individual studies have been previously published (see e.g.^40,41^) but have not been related to register-based information. Permission to link the clinical data with register data was obtained from the Danish Data Protection Agency (CSU-FCFS-2017-012, I-Suite nr: 05787). The clinical studies were approved by the Danish National committee on Biomedical Research Ethics (PECANS I: H-D-2008-088, PECANS II: H-3-2013-149, ATT: H-C-2008-076, DBUP: H-6-2014-068). All participants provided written informed consent, including parental consent for individuals <18 years.

### Participants

We included individuals with both register- and cognitive data available resulting in a final sample size of 276, including 44 children and adolescents with FEP and 49 matched HC (age 12-17), and 91 adults with FEP and 92 HC (age 18-45). All patients in the adult cohorts were antipsychotic-naïve, while approx. 65% of the child- and adolescent sample received antipsychotic medication^40,42^. Exclusion criteria for both FEP patients and HC included: a current diagnosis of drug dependence, serious somatic or neurological illness and a history of severe head injury. Exclusion criteria specific to the HC were: previous or current psychiatric illness or family history of psychiatric illness in first degree relatives.

### Assessments

Diagnoses were based on ICD-10 criteria using the Schedules for Clinical Assessment in Neuropsychiatry (SCAN)^43^ or the Schedule for Affective Disorders and Schizophrenia for School-Age Children Present and Lifetime version (K-SADS-PL)^44^. Severity of psychopathology was rated using the Positive and Negative Syndrome Scale (PANSS)^45^.

#### Cognition

Cognitive domains relevant to schizophrenia were assessed in all participants using the Brief Assessment of Cognition in Schizophrenia (BACS)^46^ covering the domains of processing speed (Symbol coding), verbal memory (List learning), working memory (Digit sequencing), motor speed (Token), planning (Tower of London) and verbal fluency (Supermarket items, F-words, and S-words). Selected tests from the Cambridge Neuropsychological Test Automated Battery (CANTAB)^47,48^ were also included to examine working memory (Spatial Span [SSP] and Spatial Working Memory [SWM]), reaction time (Reaction Time [RTI]), mental flexibility (Intra-Extra Dimensional set shift task [IED]) and sustained attention (Rapid Visual Information Processing [RVP]). Finally, IQ was estimated using four subtests (Vocabulary, Similarities, Block Design and Matrix Reasoning) from the Wechsler child or adult intelligence scales^49,50^. These subtests were selected based on their high correlation with full-scale IQ^51^.

#### Early risk factors

From the Danish Medical Birth Registry^52^, we obtained information on birth length and weight^16,53^, gestational age, winter/spring birth (December-May)^54^, paternal age^55^, maternal age, and Apgar scores^56^. The Apgar score is a rapid test routinely performed one and five minutes after birth, covering assessments of heart rate, activity (muscle tone), appearance (skin colour), grimace (reflex irritability), and breathing. Each domain is scored from 0-2, with higher scores indicating better functioning resulting in a total score of maximum 10. A total score from 4–6 is defined as abnormal, while 0–3 is defined as low^57^. Being small for the gestational age (SGA) was defined as a birth weight <10^th^ percentile using Norwegian norms for males and females separately^58^.

### Statistical analyses

Statistical analyses were performed using R (version 4.5.2). Normality was assessed by visual inspection of the histograms and the Shapiro–Wilk test. Some cognitive variables were highly skewed (From BACS: Tower of London. From CANTAB: SWM strategy and between errors, IED total errors adjusted and RVP A′), and in these cases categorical variables were created based on percentiles. Due to low variation in Apgar score we created a categorical variable: Apgar <10 yes/no. Group differences were examined using independent t-test, chi-squared test, or Mann-Whitney U-test as appropriate and FDR-corrected for multiple comparisons. The variance inflation factor (VIF) was used to examine multicollinearity. All cognitive variables had a low amount of missing data (<4%). For the risk factors, all variables were below 6.5% except for paternal age (22.1%). Missing data was imputed using multiple imputation by chained equations (MICE)^59^. 10 imputed datasets were generated with 10 iterations. Continuous variables were imputed using predictive mean matching, binary variables using logistic regression, and ordinal variables using proportional odds logistic regression. Imputation was conducted across all variables including group, sex, and age as predictors. Convergence and plausibility of imputations were assessed using trace plots and comparisons of observed and imputed variable distributions.

Canonical correlation analysis (CCA) was used to examine associations between early risk factors (X block) and cognitive domains (Y block) by identifying linear combinations of each set that were maximally correlated. Analyses were performed using the CCA package in R^60^ and performed separately within each imputed dataset. All variables were z-score standardized within each imputation before CCA. Statistical significance was assessed using permutation testing. Within each imputed dataset, rows of the cognitive variable matrix were randomly permuted (5,000 permutations) and CCA was refitted for each permutation to generate an empirical null distribution. Empirical p-values were computed as the proportion of permuted canonical correlations exceeding the observed value. Results were summarized descriptively across imputations using median and range.

Canonical loadings (structure coefficients) were used to aid interpretation. Pooled loadings were obtained by averaging sign-aligned loadings across imputations and interpreted descriptively. Uncertainty in the canonical loadings was assessed using bootstrap resampling within each imputed dataset (1,000 resamples). Participants were resampled with replacement, CCA was refitted, and canonical variates were sign-aligned to the original solution prior to computing loadings. For each variable, bootstrap means and percentile-based 95% confidence intervals were estimated within imputed datasets. Confidence intervals were conservatively combined across imputations by retaining the most extreme bounds.

We post hoc explored group differences according to diagnosis, age (<18 vs ≥18 years old), and sex. Following identification of multivariate associations via CCA, we assessed whether groups differed in their position along the canonical dimensions. To ensure comparability across imputations, canonical scores were sign-aligned to a reference imputation. Effect sizes were reported as Cohen’s *d*. Results were summarized descriptively across imputations using the median and range of permutation p-values and effect sizes.

To evaluate out-of-sample generalizability, repeated split-sample cross-validation was performed. For each imputation, the data was randomly split 10 times into training (70%) and test (30%) sets. Test-set canonical correlations were used as the primary performance metric. To assess statistical significance, permutation testing was performed by refitting CCA models on training data with permuted outcomes and evaluating the resulting test-set correlations. A pooled permutation test compared all observed test-set correlations across imputations and splits to a global empirical null distribution. Structural stability was quantified as the correlation between canonical variable loadings estimated in the training data and those obtained when projecting the held⍰out test data onto the trained canonical variates.

## RESULTS

Demographic and clinical information for HC and patients (split on child/adolescent- or adult-onset cases) are presented in **Table 1**. As expected, there were no significant differences in age or sex between patients and matched HCs. The distribution of diagnoses differed between adult and child onset patients, with a higher percentage of schizophrenia patients in the adult group. Moreover, the child FEP group had higher mean negative symptoms on the PANSS compared to adult FEP patients. Group differences in early risk factors and cognition are presented in **Table 2**. After correction for multiple comparisons, there were no differences between FEP patients and HC in the included early risk factors. FEP patients performed worse than HC on all cognitive measures except for latency measures from MOT and RVP. All VIF values were <5 indicating that multicollinearity was not a concern (Supplementary **Table S1**).

**Table 1:** Demographics and clinical information.

|  | Adult HC<br>(N=92) | Adult FEP<br>(N=91) | Child HC<br>(N=49) | Child FEP<br>(N=44) | HC vs FEP | Child vs adult FEP |
| --- | --- | --- | --- | --- | --- | --- |
| Age, mean (SD) | 23.9 (4.8) | 23.8 (4.8) | 15.9 (1.4) | 16.3 (1.2) | $t(274) = -0.3$,<br>$p = .764$ | $t(229) = 20.7$,<br>$p < .001$ |
| Sex, N (%) |  |  |  |  |  |  |
| Female | 39 (42%) | 40 (44%) | 32 (65%) | 33 (75%) | $\chi^2(1) = 0.4$,<br>$p = .536$ | $\chi^2(1) = 17.7$,<br>$p < .001$ |
| Male | 53 (58%) | 51 (56%) | 17(35%) | 11 (25%) |  |  |
| Chlorpromazine<br>equivalent (mg pr. day) | - | Ap-naïve | - | 186.6<br>(206.3) | - | - |
| ICD-10 Diagnosis, N (%) |  |  |  |  |  |  |
| F20 | - | 86 (94.5%) | - | 31 (70.5%) | - | $\chi^2(2) = 16.0$,<br>$p < .001$ |
| F25 |  | 3 (3.3%) |  | 5 (11.3%) |  |  |
| F28 |  | 2 (2.2%) |  | 8 (18.2%) |  |  |
| PANSS, mean (SD) |  |  |  |  |  |  |
| Positive | - | 18.8 (4.2) | - | 18.98 (3.3) | - | $t(133)=-0.3$, $p = .774$ |
| Negative | | 19.3 (6.8) | | 23.36 (6.3) | | $t(133)=-0.3$, $p < .001$ |
| General | | 38.5 (8.6) | | 38.07 (6.4) | | $t(133)=0.3$, $p = .754$ |
| Total | | 76.6 (16.3) | | 80.41 (11.8) | | $t(133)=-1.4$, $p = .171$ |

Note: PANSS: Positive and Negative Syndrome Scale. AP: Antipsychotics. HC: Healthy controls, FEP: First-episode psychosis. Significant effects displayed in bold.

**Table 2:** Mean values and frequency of early risk factors and cognition between groups.

|  | Adult HC | Adult FEP | Child HC | Child FEP | HC vs FEP | FDR corrected<br>p-value |
| --- | --- | --- | --- | --- | --- | --- |
| **Early risk factors** |  |  |  |  |  |  |
| Mother age, years<br>Mean (SD) | 29.1<br>(5.1) | 27.7<br>(5.5) | 30.7<br>(4.1) | 29.5<br>(5.4) | $t(266) = 2.1$,<br>$p = .034$ | $p = .138$ |
| Father age, years<br>Mean (SD) | 32.1<br>(6.2) | 31.5<br>(6.4) | 33.1<br>(5.2) | 32.5<br>(6.3) | $W = 6224.5$,<br>$p = .304$ | $p = .405$ |
| Gestational weeks<br>Mean (SD) | 39.8<br>(1.6) | 39.5<br>(1.9) | 40.2<br>(1.6) | 39.4<br>(2.1) | $W = 9523.0$,<br>$p = .069$ | $p = .183$ |
| Birth length, cm<br>Mean (SD) | 52.1<br>(2.2) | 51.6<br>(2.7) | 52.3<br>(2.0) | 51.2<br>(2.9) | $W = 10142.5$,<br>$p = .131$ | $p = .210$ |
| Birth weight, g<br>Mean (SD) | 3512.4<br>(524.3) | 3428.8<br>(617.9) | 3625.2<br>(508.7) | 3341.6<br>(585.8) | $t(263) = 2.2$,<br>$p = .029$ | $p = .138$ |
| Apgar score below 10 |  |  |  |  |  |  |
| Yes, N (%) | 6 (7.1%) | (< 5%)<sup>a</sup> | 5 (10.6%) | (< 10%)<sup>a</sup> | $\chi^2(1) = 0.4$,<br>$p = .506$ | $p = .506$ |
| No, N (%) | 78 (92.9%) | (> 95%) | 42 (89.4%) | (> 90%) |  |  |
| SGA |  |  |  |  |  |  |
| Yes, N (%) | 11 (13%) | 16 (19%) | (< 10%)<sup>a</sup> | 9 (79.5%) | $\chi^2(1) = 2.7$,<br>$p = .104$ | $p = .207$ |
| No, N (%) | 73 (87%) | 68 (81%) | (> 90%) | 35 (20.5%) |  |  |
| Winter/spring birth |  |  |  |  |  |  |
| Yes, N (%) | 43 (47%) | 51 (56%) | 24 (49%) | 21 (48%) | $\chi^2(1) = 0.7$,<br>$p = .398$ | $p = .455$ |
| No, N (%) | 49 (53%) | 40 (44%) | 25 (51%) | 23 (52%) |  |  |
| **Cognitive function** |  |  |  |  |  |  |

| Mean (SD) |  |  |  |  |  |  |
| --- | --- | --- | --- | --- | --- | --- |
| Estimated IQ | 111.8<br>(11.4) | 100.7<br>(17.0) | 107.2<br>(14.1) | 99.3<br>(19.7) | W = 11894.5,<br>p < .001 | **p < .001** |
| BACS Symbol coding | 66.6<br>(10.9) | 56.6<br>(12.8) | 58.0<br>(8.5) | 54.3<br>(10.7) | W = 12698.0,<br>p < .001 | **p < .001** |
| BACS Verbal memory | 57.1<br>(8.6) | 52.0<br>(9.7) | 52.5<br>(6.7) | 48.84<br>(8.62) | W = 11792.50,<br>p < 0.001 | **p < .001** |
| BACS Digit sequencing | 22.7<br>(3.6) | 20.6<br>(4.5) | 20.0<br>(3.5) | 17.1<br>(4.0) | W = 11982.0,<br>p < .001 | **p < .001** |
| BACS Token | 73.9<br>(13.4) | 64.1<br>(14.5) | 70.0<br>(10.4) | 61.1<br>(12.0) | W = 12392.0,<br>p < .001 | **p < .001** |
| BACS Tower of London | 19.7<br>(1.8) | 18.7<br>(2.2) | 18.8<br>(2.2) | 18.1<br>(2.2) | W = 10850.5,<br>p = .001 | **p = .001** |
| BACS Verbal fluency | 65.9<br>(12.8) | 52.6<br>(14.3) | 55.3<br>(10.7) | 46.0<br>(12.1) | t(267) = 7.2,<br>p < .001 | **p < .001** |
| MOT latency | 842.3<br>(191.3) | 893.4<br>(238.0) | 710.7<br>(137.0) | 705.3<br>(159.0) | W = 8833.0,<br>p = .469 | p = .469 |
| SSP span length | 7.7<br>(1.1) | 7.2<br>(1.3) | 7.3<br>(1.1) | 6.7<br>(1.4) | W = 11414.0,<br>p < .001 | **p = .001** |
| SWM strategy | 23.6<br>(5.0) | 28.3<br>(6.2) | 27.9<br>(5.8) | 29.9<br>(5.4) | W = 5978.0,<br>p < .001 | **p < .001** |
| SWM between errors | 5.1<br>(6.7) | 14.4<br>(18.3) | 11.1<br>(9.7) | 18.1<br>(16.8) | W = 6508.5,<br>p < .001 | **p < .001** |
| IED total errors (adj) | 16.1<br>(15.5) | 22.8<br>(19.8) | 19.0<br>(15.4) | 27.0 (19.8) | W = 6916.5,<br>p < .001 | **p < .001** |
| RTI simple motor time | 396.6<br>(135.4) | 435.2<br>(160.8) | 311.0<br>(91.0) | 364.2<br>(106.4) | W = 6893.0,<br>p = .001 | **p = .002** |
| RTI simple reaction time | 294.9<br>(42.41) | 322.78<br>(77.21) | 282.04<br>(38.50) | 318.03<br>(72.00) | W = 6550.5,<br>p < .001 | **p < .001** |
| RTI five-choice motor time | 379.0<br>(119.5) | 419.6<br>(150.2) | 301.8<br>(64.5) | 381.7<br>(240.4) | W = 6801.0,<br>p < .001 | **p = .001** |
| RTI five-choice reaction time | 321.8<br>(46.0) | 349.6<br>(63.9) | 319.9<br>(48.4) | 355.3<br>(77.0) | W = 6214.5,<br>p < .001 | **p < .001** |
| RVP A' | 0.97<br>(0.03) | 0.95<br>(0.05) | 0.89<br>(0.05) | 0.85<br>(0.06) | W = 10886.0,<br>p < .001 | **p < .001** |
| RVP latency | 349.7<br>(68.6) | 372.7<br>(119.3) | 437.9<br>(91.3) | 451.3<br>(95.8) | W = 8126.0,<br>p = .323 | p = .342 |

*Note:* Significant group difference between patients and controls after correction displayed in bold. <sup>a</sup>N and precise percentage not shown due to small cell size. BACS: Brief Assessment of Cognition in Schizophrenia. From the Cambridge Neuropsychological Test Automated Battery (CANTAB): MOT: Motor Task, SSP: Spatial Span, SWM: Spatial Working Memory, IED: Intra-Extra Dimensional set shift, RTI: Reaction Time, RVP: Rapid Visual Information Processing. HC: Healthy controls, FEP: First-episode psychosis. For the following variables, lower scores equal better performance: MOT latency, SWM strategy and between errors, IED total errors (adj.), all RTI measures and RVP latency.

### Associations between early risk factors and cognitive domains

The CCA analysis resulted in eight dimensions. CCA1 was significant in 10/10 imputations according to Wilks’ Lamda, while CCA2 was significant in 7/10. The remaining six dimensions were non-significant across imputations. Permutation testing further provided evidence that CCA1 and CCA2 exceeded that expected under the null hypothesis. Across imputations, the median empirical p-value for CCA1 was *p* = .036 (*p*_min_= .017, *p*_max_=.055) and for CCA2 it was *p* = .033 (*p*_min_= .003, *p*_max_=.168) (See Supplementary **Table S2** for permutation results from all imputations). The mean correlation between early risk factors and cognition for CCA1 was r = 0.44 [0.43;0.45] and for CCA2 it was r= 0.38 [0.36;0.40].

**Figure 1** illustrates the multivariate associations between early risk factors and cognitive domains for the first two canonical dimensions showing the average loadings across imputations. CCA1 was mainly driven by maternal age and birth length from the risk factor side and estimated IQ from the cognitive side (all loadings >0.5). Smaller but potentially meaningful contributions (loadings >0.3) were observed for SGA and birth weight to the early risk factor variate as well as working memory (both verbal and spatial), mental flexibility, processing speed, verbal fluency and motor latency to the cognitive variate. CCA2 showed potential meaningful loadings for SGA, gestational age and father age on the early risk factor side (loadings >0.3), while no loadings on the cognitive side were >0.3. For CCA1 the loadings were relatively stable across imputations, while the loadings in CCA2 were more unstable (Supplementary **Figure S1-S4**). **Figure 2** shows the bootstrapped canonical loadings.

**Figure 1:**
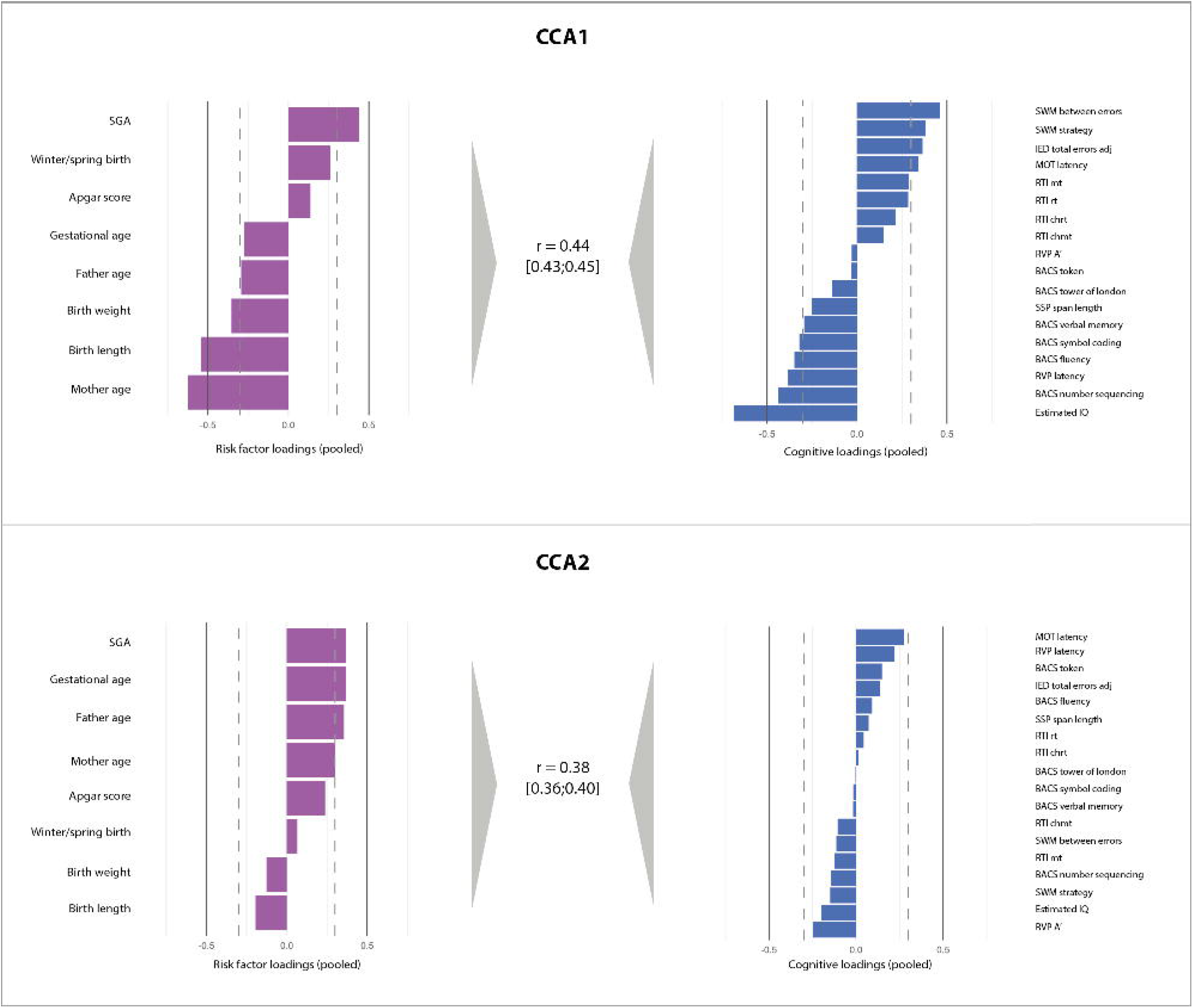
Pooled canonical loadings *Note:* BACS: Brief Assessment of Cognition in Schizophrenia. From the Cambridge Neuropsychological Test Automated Battery (CANTAB): MOT: Motor Task, SSP: Spatial Span, SWM: Spatial Working Memory, IED: Intra-Extra Dimensional set shift, RTI: Reaction Time, RVP: Rapid Visual Information Processing. For the early risk factors, a positive loading for SGA indicates that being small for the gestational age is contributing to the association. For birth weight and length as well as maternal age, negative loadings represent lower values. For the following cognitive variables, positive loadings represent poorer performance: MOT latency, all RTI measures, SWM strategy and between errors, IED total errors (adj.) and RVP latency.

**Figure 2:**
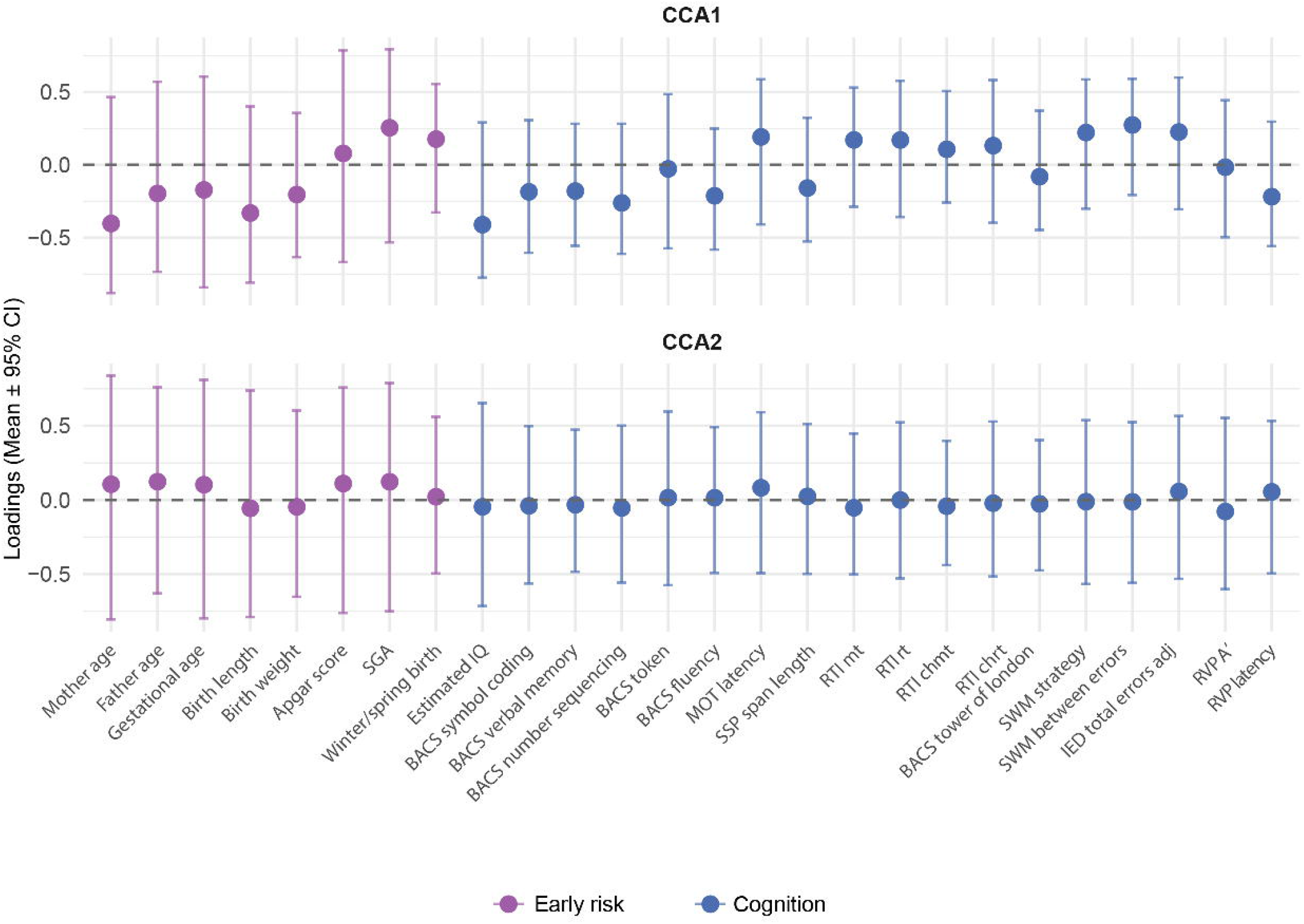
Bootstrapped canonical loadings (mean ± 95% CI) *Note*: The figure shows the bootstrapped canonical loadings for the first two canonical variates. Points represent mean loadings across bootstrap samples and imputations. Error bars indicate bootstrap variability (± 95% CI) providing an index of loading stability.

### Effects of diagnosis, age and sex

Post hoc analyses were conducted for CCA1 only. These indicated group differences according to diagnosis for both the early risk factors scores and cognitive scores. For early risk factors, permutation testing yielded a median empirical *p*-value = .004 (range: .001-.017), with a median effect size of *d* = 0.36 and for cognition we observed a median empirical *p*-value <.001 (*p*_min_<.001, *p*_max_ <.001), and a median effect size of *d* = 0.60. Moreover, we observed differences in the early risk factor scores according to age with a median empirical *p*-value = .033 (*p*_min_= .009, *p*_max_= .183) and a median effect size of *d* = 0.26. There were no group differences along CCA1 regarding sex.

### Cross-validation

Across 100 cross⍰validation evaluations (10 imputations × 10 splits), CCA1 showed a median train⍰set correlation of r = 0.50 and a median test⍰set correlation of r = 0.10 (IQR = 0.13; MAD = 0.07). CCA2 showed a median train⍰set correlation of r = 0.44 and a median test⍰set correlation of r = 0.07 (IQR = 0.12; MAD = 0.06). CCA1 showed a median X-block stability of 0.97 and Y-block stability of 0.93, with a median overall stability of 0.94 (IQR = 0.04). CCA2 showed a median X-block stability of 0.95 and Y-block stability of 0.87, with a median overall stability of 0.90 (IQR = 0.07). Pooled permutation testing of the test-set correlations using a global empirical null distribution yielded p⍰values of 0.46 for CCA1 and 0.56 for CCA2.

## DISCUSSION

The aim of the study was to examine multivariate patterns of associations between early risk factors for schizophrenia obtained at birth and a broad range of cognitive domains assessed many years later at the time of a first-episode psychosis diagnosis. Our findings revealed two meaningful multivariate associations. The first canonical dimension (CCA1) was stable across imputations and the significance was supported by permutation testing, indicating a robust association. However, the bootstrap-derived confidence intervals for the individual loadings were wide and crossing zero, indicating uncertainty in the precise contribution of specific variables. This could reflect sampling variability and bias introduced by multiple imputations rather than an absence of a multivariate effect. Examination of the loadings revealed that lower maternal age, lower birth length, being small for gestational age and lower birth weight constituted an early risk profile. For cognition, lower estimated IQ, poorer processing speed, working memory, verbal fluency and mental flexibility showed meaningful contributions to the association. This is consistent with previous findings showing a positive association between birth weight and cognitive functioning in both non-psychiatric populations^17–20^ and in patients with schizophrenia^35,36^ as well as an association between being small for gestational age and lower IQ^19,21,22^. Lower maternal age also contributed to the multivariate association consistent with a previous meta-analysis showing an association between low maternal age and more cognitive problems in the offspring^26,61^. One could speculate that this finding is related to being primigravida. Finally, poorer working memory (both verbal and spatial), mental flexibility, verbal fluency and motor speed also showed meaningful contributions to the cognitive side, hereby extending the previous literature showing that early risk factors influence not only IQ but cognitive performance more broadly later in life.

For the second canonical dimension (CCA2), permutation testing supported a relationship beyond chance, however the significance of CCA2 was more unstable across imputations. Furthermore, the loadings were relatively unstable across imputations, and thus we abstain from interpreting the specific variables contributing to the profile of CCA2.

We did not observe any meaningful contributions from several early risk factors (i.e., advanced paternal age, winter/spring birth and Apgar scores). Although a relationship between paternal age and IQ in the offspring has been reported previously^24^, another study suggested that this association may be explained by other factors, e.g. maternal age^25^, which would be consistent with our findings. However, the lack of an association in our study could also be explained by the data quality of this variable, as this was the variable with the largest amount of missing data (<20%). We did not find an association between winter birth and cognition in these analyses in line with some previous studies^31^, although findings are mixed^28,29,32^ and may also be explained by other factors such as maternal IQ^30^, underscoring the importance of a multivariate approach. Finally, in this cohort the majority (>90% across groups) had an Apgar score of 10 (highest score). The lack of variation in this measure may explain why we did not observe any meaningful associations with cognition.

As an exploratory aim, we examined potential group differences according to diagnosis, sex, and age group. Post hoc analyses showed group differences along the first canonical dimension (CCA1) according to diagnosis, indicating that patients showed a greater expression of the multivariate pattern compared to HCs. This was the case for both early risk factors and cognition with moderate effect sizes, reflecting greater overall exposure to the early risk factor profile as well as the poorer cognitive performance profile in patients. Moreover, adults (as a whole group including both FEP and HCs) showed greater expression of the early risk canonical scores compared to children and adolescents, however the effect size was small.

Finally, we evaluated the out-of-sample generalizability using repeated cross-validation. Both CCA1 and CCA2 showed substantially lower correlations in held-out test data compared with training data, indicating limited generalization. Here it is important to note that canonical correlations are by design maximized in sample, and cross-validation disrupts this optimization and some shrinkage between train- and test-set correlations is expected. Our test-set correlations were of similar magnitude to a previous study^62^, however, permutation testing indicated that the out-of-sample correlations did not exceed chance levels, underscoring the need for replication in independent samples. Nevertheless, structural stability was high for both CCA1 and CCA2, suggesting that the patterns were reproducible across training and test samples despite attenuation in canonical correlation magnitude.

The current findings should be considered within the strengths and limitations of the study. We included Danish register-based information collected at the time of birth on the early risk factors to minimize recall bias. Moreover, the combination of register-based information with detailed clinical data is unique and pose a substantial strength. We assessed the stability of our findings across imputations and the robustness using permutation testing and bootstrapping according to proposed guidelines for CCA^63^. Moreover, we evaluated the out-of-sample generalizability using cross-validation. Nevertheless, CCA has several inherent limitations including being susceptible to overfitting and sample-specific solutions. Moreover, CCA assumes linear relationships and may not capture more complex, non-linear associations between early risk factors and cognitive outcomes.

We included a relatively large sample of patients with first-episode psychosis covering a broad age range. However, the child- and adolescent sample was smaller, which is a limitation. Moreover, the imputation of missing data on some variables may have introduced unknown bias. While the adult patients were completely antipsychotic-naïve at the time of the cognitive assessment, the child/adolescent sample had received some treatment with antipsychotic medication, which could be considered a limitation. Nevertheless, in a previous study using the same cohort, medication dose was only associated with a few cognitive measures, i.e. planning and reaction time^40^.

In conclusion, multiple early risk factors obtained at birth are associated with poorer cognitive functioning across different domains later in life, indicating that no single risk factor drives cognitive outcomes in isolation. This underscores the complexity of developmental processes and the importance of the early context in shaping cognitive trajectories. Particularly lower maternal age, lower birth length and weight and being small for the gestational age contributed to a cognitive profile of lower estimated IQ, poorer working memory, mental flexibility, processing speed and verbal fluency. These risk factors are routinely collected and could easily be incorporated into clinical practice for risk-based stratification to support early identification and intervention, ultimately improving cognitive outcomes.

## Data Availability

All data produced in the present study are available upon reasonable request to the authors and following legal acceptance

## ACKNOWLEDGEMENTS AND FUNDING

The authors would like to thank all the patients and controls for participating in the studies. Moreover, we would like to thank the participating mental health centres and all the researchers working on the clinical studies over the years for carrying out cognitive and clinical assessments.

The current study was funded by the Mental Health Services in the Capital Region of Denmark and the Lundbeck Foundation (grant numbers: R25-A2701 and R155-2013-16337). The child- and adolescent cohorts were further supported by Læge Gerhard Linds Legat; Fru C. Hermansens Legat; Slagtermester Wörzners og Hustru Inger Wörzners Mindelegat; Psykiatrisk Forskningsfond af 1967; Tømrermester Jørgen Holm og Hustru Elisa F. Hansens Mindelegat and Jascha Fonden. No funding source were involved in the development or interpretation of the study.

CP was supported by an Australian National Health and Medical Research Council (NHMRC) L3 Investigator Grant (1196508).

## REFERENCES

1. Catalan A, McCutcheon RA, Aymerich C, et al. The magnitude and variability of neurocognitive performance in first-episode psychosis: a systematic review and meta-analysis of longitudinal studies. Transl Psychiatry. Springer Nature. 2024;14(1). doi:10.1038/s41398-023-02718-6

2. Lee M, Cernvall M, Borg J, et al. Cognitive Function and Variability in Antipsychotic Drug-Naive Patients with First-Episode Psychosis: A Systematic Review and Meta-Analysis. JAMA Psychiatry. 2024;81(5):468–476. doi:10.1001/jamapsychiatry.2024.0016

3. Murray RM, Bhavsar V, Tripoli G, Howes O. 30 Years on: How the Neurodevelopmental Hypothesis of Schizophrenia Morphed into the Developmental Risk Factor Model of Psychosis. Schizophr Bull. 2017;43(6):1190–1196. doi:10.1093/schbul/sbx121

4. Mollon J, David AS, Zammit S, Lewis G, Reichenberg A. Course of cognitive development from infancy to early adulthood in the psychosis spectrum. JAMA Psychiatry. 2018;75(3):270–279. doi:10.1001/jamapsychiatry.2017.4327

5. Tandon R, Nasrallah HA, Keshavan MS. Schizophrenia, “just the facts” 4. Clinical features and conceptualization. Schizophr Res. 2009;110(1-3):1–23. doi:10.1016/j.schres.2009.03.005

6. Pedersen CB, Mors O, Bertelsen A, et al. A comprehensive nationwide study of the incidence rate and lifetime risk for treated mental disorders. JAMA Psychiatry. 2014;71(5):573–581. doi:10.1001/jamapsychiatry.2014.16

7. Werry JS. Child and Adolescent (Early Onset) Schizophrenia: A review in light of DSM-III-R. J Autism Dev Disord. 1992;22(4):601–624. doi:10.1007/BF01046330

8. Salazar De Pablo G, Rodriguez V, Besana F, et al. Umbrella Review: Atlas of the Meta-Analytical Evidence of Early-Onset Psychosis. Vol 63. 2024. www.jaacap.org

9. Smelror RE, Johannessen C, Wedervang-Resell K, et al. Cognitive impairment profile in adolescent early-onset psychosis using the MATRICS Battery: Age and sex effects. Neuropsychology. 2021;35(3):300–309. doi:10.1037/neu0000723

10. Blokland GAM, Mesholam-Gately RI, Toulopoulou T, et al. Heritability of Neuropsychological Measures in Schizophrenia and Nonpsychiatric Populations: A Systematic Review and Meta-analysis. Schizophr Bull. Oxford University Press. 2017;43(4):788–800. doi:10.1093/schbul/sbw146

11. Lemvigh C, Brouwer R, Sahakian B, et al. Heritability of Memory Functions and Related Brain Volumes: A Schizophrenia Spectrum Study of 214 Twins. Schizophr Bull Open. 2020;1(1):1–11.

12. Lemvigh C, Brouwer R, Pantelis C, et al. Heritability of Specific Cognitive Functions and Associations with Schizophrenia Spectrum Disorders using CANTAB: A Nation-wide Twin study. Psychol Med. Published online 2020:1–14. doi:10.1017/S0033291720002858

13. Hilker R, Helenius D, Fagerlund B, et al. Heritability of Schizophrenia and Schizophrenia Spectrum Based on the Nationwide Danish Twin Register. Biol Psychiatry. 2018;83(6):492–498. doi:10.1016/j.biopsych.2017.08.017

14. Owens SF, Rijsdijk F, Picchioni MM, et al. Genetic overlap between schizophrenia and selective components of executive function. Schizophr Res. 2011;127(1-3):181–187. doi:10.1016/j.schres.2010.10.010

15. Lemvigh CK, Ambrosen KS, Ebdrup BH, Glenthøj BY, Osler M, Fagerlund B. Impact of early risk factors on schizophrenia risk and age of diagnosis: A Danish population-based register study. European Psychiatry. 2024;67(1):e64. doi:10.1192/j.eurpsy.2024.1774

16. Radua J, Ramella-Cravaro V, Ioannidis JPA, et al. What causes psychosis? An umbrella review of risk and protective factors. World Psychiatry. 2018;17(1):49–66. doi:10.1002/wps.20490

17. Eves R, Mendonça M, Baumann N, et al. Association of Very Preterm Birth or Very Low Birth Weight with Intelligence in Adulthood: An Individual Participant Data Meta-analysis. JAMA Pediatr. 2021;175(8). doi:10.1001/jamapediatrics.2021.1058

18. Kormos CE, Wilkinson AJ, Davey CJ, Cunningham AJ. Low birth weight and intelligence in adolescence and early adulthood: A meta-analysis. Journal of Public Health (United Kingdom). 2014;36(2):213–224. doi:10.1093/pubmed/fdt071

19. Sania A, Sudfeld CR, Danaei G, et al. Early life risk factors of motor, cognitive and language development: A pooled analysis of studies from low/middle-income countries. BMJ Open. 2019;9(10). doi:10.1136/bmjopen-2018-026449

20. Flensborg-Madsen T, Mortensen EL. Birth weight and intelligence in young adulthood and midlife. Pediatrics. 2017;139(6). doi:10.1542/peds.2016-3161

21. Gonçalves R, Blaauwendraad S, Avraam D, et al. Early-Life Growth and Emotional, Behavior and Cognitive Outcomes in Childhood and Adolescence in the EU Child Cohort Network: Individual Participant Data Meta-Analysis of over 109,000 Individuals. Vol 52. 2025. www.thelancet.com

22. Nguyen PT, Nguyen PH, Tran LM, et al. The Relationship of Preterm and Small for Gestational Age with Child Cognition During School-Age Years. Journal of Nutrition. 2024;154(8):2590–2598. doi:10.1016/j.tjnut.2024.06.012

23. Husby A, Wohlfahrt J, Melbye M. Gestational age at birth and cognitive outcomes in adolescence: population based full sibling cohort study. BMJ. Published online 2023. doi:10.1136/bmj-2022-072779

24. Zweifel JE, Woodward JT. The risky business of advanced paternal age: neurodevelopmental and psychosocial implications for children of older fathers. Fertil Steril. Elsevier Inc. 2022;118(6):1013–1021. doi:10.1016/j.fertnstert.2022.10.029

25. McGrath J, Mortensen PB, Pedersen CB, Ehrenstein V, Petersen L. Paternal Age and General Cognitive Ability-A Cross Sectional Study of Danish Male Conscripts. PLoS One. 2013;8(10). doi:10.1371/journal.pone.0077444

26. Cresswell L, Faltyn M, Lawrence C, et al. Cognitive and Mental Health of Young Mothers’ Offspring: A Meta-analysis. Pediatrics. American Academy of Pediatrics. 2022;150(5). doi:10.1542/peds.2022-057561

27. Goisis A, Schneider DC, Myrskylä M. The reversing association between advanced maternal age and child cognitive ability: Evidence from three UK birth cohorts. Int J Epidemiol. 2017;46(3):850–859. doi:10.1093/ije/dyw354

28. McGrath JJ, Saha S, Lieberman DE, Buka S. Season of birth is associated with anthropometric and neurocognitive outcomes during infancy and childhood in a general population birth cohort. Schizophr Res. 2006;81(1):91–100. doi:10.1016/j.schres.2005.07.017

29. Bai Y, Shang G, Wang L, Sun Y, Osborn A, Rozelle S. The relationship between birth season and early childhood development: Evidence from northwest rural China. PLoS One. 2018;13(10). doi:10.1371/journal.pone.0205281

30. Grootendorst-van Mil NH, M Steegers-Theunissen RP, Hofman A, V Jaddoe VW, Verhulst FC, Tiemeier H. Brighter children? The association between seasonality of birth and child IQ in a population-based birth cohort. BMJ Open. 2017;7. doi:10.1136/bmjopen-2016

31. Hori H, Teraishi T, Sasayama D, et al. Relationships between season of birth, schizotypy, temperament, character and neurocognition in a non-clinical population. Psychiatry Res. 2012;195(1-2):69–75. doi:10.1016/j.psychres.2011.07.028

32. Al-Sabah R, Al-Taiar A, Rahman A, Shaban L, Al-Harbi A, Mojiminiyi O. Season of birth and sugary beverages are predictors of Raven’s Standard Progressive Matrices Scores in adolescents. Sci Rep. 2020;10(1). doi:10.1038/s41598-020-63089-2

33. Amoretti S, Rabelo-Da-Ponte FD, Garriga M, et al. Obstetric complications and cognition in schizophrenia: A systematic review and meta-analysis. Psychol Med. Cambridge University Press. 2022;52(14):2874–2884. doi:10.1017/S0033291722002409

34. Teigset CM, Mohn C, Rund BR. Perinatal complications and executive dysfunction in early-onset schizophrenia. BMC Psychiatry. 2020;20(1). doi:10.1186/s12888-020-02517-z

35. Freedman D, Bao Y, Kremen WS, Vinogradov S, McKeague IW, Brown AS. Birth weight and neurocognition in schizophrenia spectrum disorders. Schizophr Bull. 2013;39(3):592–600. doi:10.1093/schbul/sbs008

36. Torniainen M, Wegelius A, Tuulio-Henriksson A, Lönnqvist J, Suvisaari J. Both low birthweight and high birthweight are associated with cognitive impairment in persons with schizophrenia and their first-degree relatives. Psychol Med. 2013;43(11):2361–2367. doi:10.1017/S0033291713000032

37. Nielsen MØ, Rostrup E, Wulff S, et al. Alterations of the brain reward system in antipsychotic nave schizophrenia patients. Biol Psychiatry. 2012;71(10):898–905. doi:10.1016/j.biopsych.2012.02.007

38. Bojesen KB, Ebdrup BH, Jessen K, et al. Treatment response after 6 and 26 weeks is related to baseline glutamate and GABA levels in antipsychotic-naïve patients with psychosis. Psychol Med. Published online September 2019:1–12. doi:10.1017/S0033291719002277

39. Rydkjær J, Møllegaard Jepsen JR, Pagsberg AK, Fagerlund B, Glenthøj BY, Oranje B. Mismatch negativity and P3a amplitude in young adolescents with first-episode psychosis: a comparison with ADHD. Psychol Med. 2017;47(02):377–388. doi:10.1017/S0033291716002518

40. Jepsen JRM, Rydkjaer J, Fagerlund B, et al. Cross-sectional associations between adaptive functioning and social cognitive and neurocognitive functions in adolescents with first-episode, early-onset schizophrenia spectrum disorders. Dev Psychopathol. Published online February 1, 2022:1–11. doi:10.1017/S0954579422001110

41. Fagerlund B, Pantelis C, Jepsen JRM, et al. Differential effects of age at illness onset on verbal memory functions in antipsychotic-naïve schizophrenia patients aged 12 – 43 years. Psychol Med. Published online 2020. doi:10.1017/S0033291720000409

42. Lemvigh C, Jepsen JRM, Fagerlund B, et al. Auditory sensory gating in young adolescents with early-onset psychosis: A comparison with ADHD. Neuropsychopharmacology. 2019;0(October):1–7. doi:10.1093/schbul/sby017.565 LK - http://AD4MH3SR7V.search.serialssolutions.com?sid=EMBASE&issn=17451701&id=doi:10.1093%2Fschbul%2Fsby017.565&atitle=Auditory+sensory+gating+in+young+adolescents+with+early-onset+psychosis%3A+A+comparison+with+ADHD&stitle=Schizophr.+Bull.&title=Schizophrenia+Bulletin&volume=44&issue=&spage=S232&epage=&aulast=Lemvigh&aufirst=Cecilie&auinit=C.&aufull=Lemvigh+C.&coden=&isbn=&pages=S232-&date=2018&auinit1=C&auinitm=

43. Wing JK, Babor T, Brugha T, et al. SCAN. Schedules for Clinical Assessment in Neuropsychiatry. Arch Gen Psychiatry. 1990;47(6):589–593.

44. Kaufman J, Birmaher B, Brent D, et al. Schedule for Affective Disorders and Schizophrenia for School-Age Children-Present and Lifetime Version (K-SADS-PL): Initial Reliability and Validity Data. J Am Acad Child Adolesc Psychiatry. 1997;36(7):980–988.

45. Kay SR, Fiszbein A, Opler LA. The Positive and Negative Syndrome Scale (PANSS) for Schizophrenia. Schizophr Bull. 1987;13(2):261–276. doi:10.1093/schbul/13.2.261

46. Keefe R, Goldberg T, Harvey P, Gold J, Poe M, Coughenour L. The Brief Assessment of Cognition in Schizophrenia: Reliability, sensitivity, and comparison with a standard neurocognitive battery. Schizophr Res. 2004;68(2-3):283–297. doi:10.1016/j.schres.2003.09.011

47. Evenden J, Morris R, Owen A, Robbins T, Roberts A, Sahakian B. CANTABeclipse Test Administration Guide. Manual ver. Cambridge Cognition Limited: Cambridge; 2013.

48. Sahakian BJ, Owen AM. Computerized assessment in neuropsychiatry using CANTAB: Discussion paper. J R Soc Med. 1992;85(7):399–402.

49. Wechsler D. Manual for the Wechsler Intelligence Scale for Children, Fourth Edition. Technical and Interpretative Manual. Psychological Corporation. Preprint posted online 2003.

50. Wechsler D. Manual for the Wechsler Adult Intelligence Scale - Third Edition (WAIS-III). The Psychological Corporation; 1997.

51. Axelrod BN. Validity of the Wechsler Abbreviated Scale of Intelligence and other very short forms of estimating intellectual functioning. Assessment. 2002;9(1):17–23. doi:10.1177/1073191102009001003

52. Bliddal M, Broe A, Pottegård A, Olsen J, Langhoff-Roos J. The Danish Medical Birth Register. Eur J Epidemiol. 2018;33(1):27–36. doi:10.1007/s10654-018-0356-1

53. Byrne M, Agerbo E, Bennedsen B, Eaton WW, Mortensen PB. Obstetric conditions and risk of first admission with schizophrenia: A Danish national register based study. Schizophr Res. 2007;97(1-3):51–59. doi:10.1016/j.schres.2007.07.018

54. Davies G, Welham J, Chant D, Torrey EF, McGrath J. A Systematic Review and Meta-analysis of Northern Hemisphere Season of Birth Studies in Schizophrenia. Schizophr Bull. 2003;29(3):587–593. doi:10.1093/oxfordjournals.schbul.a007030

55. Torrey EF, Buka S, Cannon TD, et al. Paternal age as a risk factor for schizophrenia: How important is it? Schizophr Res. 2009;114(1-3):1–5. doi:10.1016/j.schres.2009.06.017

56. Watterberg KL, Aucott S, Benitz WE, et al. The apgar score. Pediatrics. 2015;136(4):819–822. doi:10.1542/peds.2015-2651

57. American College of Obstetricians and Gynecologists. The Apgar Score. Committee Opinion No 644. Obstetrics & Gynecology. 2015;126(644):52–55.

58. Skjærven R, Gjessing HK, Bakketeig LS. Birthweight by gestational age in Norway. Acta Obstet Gynecol Scand. 2000;79(6):440–449. doi:10.1034/j.1600-0412.2000.079006440.x

59. Van Buuren S, Groothuis-Oudshoorn K. mice: Multivariate Imputation by Chained Equations in R. J Stat Softw. 2011;45(3). http://www.jstatsoft.org/

60. González I& DS. CCA: Canonical Correlation Analysis. Preprint posted online 2023.

61. Falster K, Hanly M, Banks E, et al. Maternal age and offspring developmental vulnerability at age five: A population-based cohort study of Australian children. PLoS Med. 2018;15(4). doi:10.1371/journal.pmed.1002558

62. Milecki L, Gonzalez C, Adeli E, et al. Regularized CCA identifies sex-specific brain-behavior associations in adolescent psychopathology. Transl Psychiatry. 2025;15(1). doi:10.1038/s41398-025-03678-9

63. Mihalik A, Chapman J, Adams RA, et al. Canonical Correlation Analysis and Partial Least Squares for Identifying Brain–Behavior Associations: A Tutorial and a Comparative Study. Biol Psychiatry Cogn Neurosci Neuroimaging. Elsevier Inc. 2022;7(11):1055–1067. doi:10.1016/j.bpsc.2022.07.012

64. Nielsen MØ, Kristensen TD, Borup Bojesen K, Glenthøj BY, Lemvigh CK, Ebdrup BH. Differential Effects of Aripiprazole and Amisulpride on Negative and Cognitive Symptoms in Patients With First-Episode Psychoses. Front Psychiatry. 2022;13. doi:10.3389/fpsyt.2022.834333

65. Feber L, Peter NL, Chiocchia V, et al. Antipsychotic Drugs and Cognitive Function: A Systematic Review and Network Meta-Analysis. JAMA Psychiatry. 2025;82(1):47–56. doi:10.1001/jamapsychiatry.2024.2890

